# Pregnancy and breastfeeding during COVID-19 pandemic: A systematic review of published pregnancy cases

**DOI:** 10.1101/2020.04.25.20079509

**Authors:** Carina Rodrigues, Inês Baía, Rosa Domingues, Henrique Barros

**Affiliations:** EPIUnit - Instituto de Saúde Pública, Universidade do Porto, Porto, Portugal; Instituto Nacional de Infectologia Evandro Chagas / Fundação Oswaldo Cruz, Rio de Janeiro, Brasil; Departamento de Ciências da Saúde Pública e Forenses e Educação Médica, Faculdade de Medicina, Universidade do Porto, Porto, Portugal

**Keywords:** COVID-19, SARS-COV-2, Pregnancy, Vertical transmission, Breastfeeding, Perinatal outcomes, systematic review

## Abstract

**Background:** The COVID-19 pandemic is an emerging concern regarding the potential adverse effects during pregnancy. This study reviews knowledge on the impact of COVID-19 on pregnancy and describes the outcome of published cases of pregnant women diagnosed with COVID-19.

**Methods:** Searches were conducted in PubMed® up to 8 April 2020, using PRISMA standards, to identify original published studies describing pregnant women at any gestational age diagnosed COVID-19. There were no date or language restrictions on the search. All identified studies were included irrespective of assumptions on study quality.

**Results:** We identified 30 original studies reporting 212 cases of pregnant women with COVID-19 (30 discharged while pregnant), 200 from China and 12 from other countries. The 182 published deliveries resulted in one stillbirth and 185 live births. Four women with severe COVID-19 required admission to an intensive care unit but no cases of maternal death were reported. There was one neonatal death. Preterm births occurred in 28.7% of cases, but it is unclear whether this was iatrogenic. All cases with amniotic fluid, placenta, and/or cord blood analyzed for the SARS-CoV-2 virus were negative. Four newborns were positive for SARS-CoV-2 and three newborns had high levels of IgM antibodies. Breast milk samples from 13 mothers and described in seven studies showed no evidence of SARS-CoV-2.

**Conclusion:** The evidence related to the effect of COVID-19 on pregnant women is still limited. Pregnant women and newborns should be considered particularly vulnerable populations regarding COVID-19 prevention and management strategies.

## Introduction

The disease resulting from infection with the Severe Acute Respiratory Syndrome Coronavirus 2 (SARS-COV-2) and designated COVID-19 by the World Health Organization (WHO) was first identified in humans in December 2019, in the city of Wuhan, China,^1^ and can present from asymptomatic to a severe acute respiratory infection requiring intensive care.^2, 3^ The infection can occur at any age, but COVID-19 is proportionally uncommon in children (<1% of the total cases). The infection fatality rate is around 1% but much higher in older people or those with pre-existing medical conditions (such as heart disease, diabetes, COPD).^2, 4^

Person-to-person transmission of COVID-19 is well established and can occur when an infected person coughs, sneezes or speaks and scattered droplets are inhaled or reach the mucous membranes of the mouth, nose or eyes of susceptible. COVID-19 can also be transmitted through direct hand contact with surfaces or objects contaminated with SARS-CoV-2 followed by contact with the mouth, nose or eyes.^2^ Pregnant women and newborns receive special attention and there is an emerging concern with the potential risk of SARS-COV-2 vertical transmission (from mother to fetus) or associated malformations, and contagion during delivery and breastfeeding; likewise, it is important to determine the potential adverse effects of COVID-19 in pregnant women.^5–8^ However, in general, the available information remains scarce.

This study reviews published cases of pregnant women diagnosed with COVID-19.

## Methods

The review follows the Preferred Reporting of Systematic Reviews and Meta-Analysis (PRISMA) guidelines.^9, 10^

We searched PubMed® up to 8 April 2020 to identify original published studies describing pregnant women at any gestational age diagnosed with COVID-19 (confirmed by clinical/radiological evidence of pneumonia compatible with SARS-CoV-2 and/or by quantitative real-time polymerase chain reaction (PCR) or dual fluorescence PCR of SARS-CoV-2 infection). The following search expression was used [(COVID-19 OR 2019-nCoV OR “novel coronavirus” OR SARS-CoV-2 OR “coronavirus 2”) AND (pregnancy OR delivery OR pregnant OR obstetric* OR maternal OR perinatal OR breastfeeding)]. Also, reference tracking was carried out to identify other potential studies to be included.

Each reference retrieved was screened independently by two researchers (IB and RD) following predefined criteria to determine eligibility for the systematic review. Studies were excluded if: (1) did not involve humans (e.g. in vitro or animal research); (2) non-original articles (e.g. book chapters, review articles, editorials, comments, guidelines); (3) data not reporting pregnant women diagnosed with COVID-19; and (4) duplicate studies or evaluating the same sample. There were no date or language restrictions on the search. Two researchers (IB and RD) reviewed the included studies and extracted the following data: collection period, maternal age, pregnancy complications, type of delivery, indication for cesarean section, gestational age at birth (or at admission), pregnancy outcome, maternal admission to intensive care unit, maternal death, neonatal outcomes (birth weight, Apgar at 1 and 5 minutes, neonatal complications, breastfeeding), intrauterine and/or neonatal samples collected for detection of SARS-CoV-2 (such as amniotic fluid, cord blood, placenta, breast milk, nasopharyngeal and anal swabs) and their results (negative/positive).

All identified original studies reporting cases of pregnant women at any gestational age diagnosed with COVID-19 were included irrespective of study quality. Cases reported in more than one study, and for which it was possible to identify duplicates, were described only once, presenting the more detailed data. We identified duplicates based on author names and hospital location, publication date, participant admission date, maternal and neonatal characteristics and outcomes.

Doubts on possible duplicates and/or differences in the data extraction were discussed and resolved by consensus, involving a third researcher (CR) whenever necessary.

## Results

Table 1 summarizes the main characteristics of the 212 reported cases of pregnant women diagnosed with COVID-19 and identified in 30 original studies published until April 8, 2020. A detailed description of each reported case is presented in Table S1 (supplemental material).

**Table 1.** Characteristics of pregnant women diagnosed with COVID-19 described in the literature (n=212)

|  | n/N (%) |
| --- | --- |
| **Maternal characteristics** |  |
| Age, years (min-max) | 22-43 |
| Country of hospital admission |  |
| China | 200/212 (94.3) |
| United States of America | 8/212 (3.8) |
| South Korea | 1/212 (0.5) |
| Honduras | 1/212 (0.5) |
| Turkey | 1/212 (0.5) |
| Sweden | 1/212 (0.5) |
| **Pregnancy complications** |  |
| Fetal distress | 20/212 (9.4) |
| Gestational diabetes | 13/212 (6.1) |
| Gestational hypertension | 11/212 (5.2) |
| PROM | 7/212 (3.3) |
| Anemia | 5/212 (2.4) |
| Placenta previa/bleeding | 5/212 (2.4) |
| Preeclampsia | 4/212 (1.9) |
| Hypothyroidism | 4/212 (1.9) |
| Chronic hypertension | 3/212 (1.4) |
| Thalassemia | 2/212 (0.9) |
| Polycystic ovary syndrome | 2/212 (0.9) |
| Cholecystitis | 2/212 (0.9) |
| Asthma | 2/212 (0.9) |
| Diabetes mellitus type 2 | 2/212 (0.9) |
| Placental abruption | 1/212 (0.5) |
| Oligohydramnios | 1/212 (0.5) |
| Polyhydramnios | 1/212 (0.5) |
| Hepatitis B infection | 1/212 (0.5) |
| Mitral valve and tricuspid valve replacement | 1/212 (0.5) |
| **Pregnancy characteristics/outcomes** |  |
| Pregnant women at discharge (undelivered) | 30/212 (14.2) |
| Multiple pregnancies | 4/212 (1.9) |
| Deliveries | 182/212 (85.8) |
| Multiple pregnancies delivered | 4/182 (2.2) |
| Stillbirths | 1/186 (0.5) |
| Live births | 185/186 (99.5) |
| Cesarean section | 139/158 (88.0) |
| Preterm birth (<37 weeks of gestation) | 45/157 (28.7) |
| **Maternal outcomes** |  |
| Maternal deaths | 0/212 (0.0) |
| Maternal admission to Intensive Care Unit | 4/212 (1.9) |
| **Neonatal outcomes** |  |
| Neonatal deaths | 1/185 (0.5) |
| SARS-CoV-2 infection confirmed by oral swabs | 4/185 (2.2) |
| High levels of SARS-CoV-2 IgM antibodies | 3/185 (1.6) |
| **Type of intrauterine/ neonatal samples collected and results** |  |
| Placenta | 21/186 (11.3) |
| Positive results | 0/21 (0.0) |
| Cord blood | 13/186 (7.0) |
| Positive results | 0/13 (0.0) |
| Amniotic fluid | 14/186 (7.5) |
| Positive results | 0/14 (0.0) |
| Newborn's oral swabs | 70/185 (33.8) |
| Positive results | 4/70 (5.7) |
| Measurement of SARS-CoV-2 IgM and IgG antibody | 7/185 (3.8) |
| Reactive IgM | 3/7 (42.8) |
| Reactive IgG | 6/7 (85.7) |
| Breast milk | 13/185 (7.0) |
| Positive results | 0/13 (0.0) |

All published cases occurred in China, except eight from the United States of America,^11,12^ one from South Korea,^13^ one from Honduras,^14^ one from Turkey,^15^ and another from Sweden.^16^ Maternal age ranged from 22 to 41 years. From the 212 pregnant women described, 182 delivered and 30 were discharged during pregnancy (undelivered). Most women were in the third trimester of pregnancy and there was only one study reporting pregnant women in the first trimester.^17^

### Vertical transmission of COVID-19

All cases in which amniotic fluid, placenta, and/or cord blood were analyzed for SARS-CoV-2 virus were negative.^5, 11, 13, 15, 18–26^

Most studies detected the SARS-CoV-2 RNA by real-time reverse transcription-polymerase chain reaction (RT-PCR) using samples from the newborn’s nasopharyngeal or throat, sample collection varying from immediately to 9 days after birth. Two studies also used maternal and neonatal sera samples to test for IgG and IgM antibodies.^27, 28^ Four newborns (2.2%) presented positive oral swabs for SARS-CoV-2.^21, 24, 25, 29^ In a cohort of 33 newborns from mothers with COVID-19, admitted to the Pediatric Hospital of Wuhan, three of the newborns had positive RT-PCR for SARS-CoV-2 in nasopharyngeal and anal swabs collected on the second and fourth days after delivery, with negative results on the sixth day for two newborns and on the seventh day for another. However, placenta, amniotic fluid, and cord blood samples were not analyzed.^29^ There is yet another case of a newborn with a positive RT-PCR for SARS COV-2 in one throat swab collected at 36 hours after birth.^21, 24^ However, it was not possible to confirm whether it was a real case of intrauterine transmission since the umbilical cord and placenta blood samples were negative for SARS-CoV-2 and the possibility of postnatal contact could not be discarded.^21, 24^ No other study reported positive results for the SARS-CoV-2 virus in nasopharyngeal exudates from newborns of mothers diagnosed with COVID-19 ^5, 11–20, 22–26, 29–36^

In a series of six cases that had blood collected after delivery evaluated, two of the newborns had high levels of IgG and IgM antibodies (>10 AU/mL) and three had high values of IgG antibodies with normal levels of IgM in, but in none SARS-CoV-2 virus was detected by RT-PCR in the oropharyngeal exudate.^27^ A case study also reported high values of IgM and IgG antibodies in the blood at days 1 and 15 after delivery, but with RT-PCR for SARS-CoV-2 negative in five samples of nasopharyngeal exudates collected between the first two hours and the 16^th^ day of life.^28^

Seven studies reported the test of breast milk samples from 13 mothers and none evidenced SARS-CoV-2 virus.^5, 15, 16, 20–22, 28^

### Maternal and neonatal outcomes

The clinical and obstetric conditions most frequently reported were fetal distress (n=20), gestational diabetes (n=13), gestational hypertension (n=11), premature rupture of membranes (PROM) (n=7), anemia (n=5), placenta previa/bleeding in the third trimester (n=5), pre-eclampsia (n=4), hypothyroidism (n=4), chronic hypertension (n=3), thalassemia(n=2), polycystic ovary syndrome (n=2), cholecystitis (n=2), asthma (n=2), diabetes mellitus type 2 (n=2), placental abruption (n=1), oligohydramnios(n=1), polyhydramnios (n=1), hepatitis B infection (n=1) and mitral valve and tricuspid valve replacement (n=1). In one study that compared groups of pregnant women with and without COVID-19, there were no significant differences in the occurrence of gestational diabetes, severe pre-eclampsia, PROM, fetal distress, meconium-stained amniotic fluid, premature delivery, neonatal asphyxia and procedures for severe post-partum bleeding.^30^ Cesarean section was the most common type of delivery: 88.0% of 158 cases with available information. Most studies did not specify the indication for the cesarean section. Four pregnant women with severe COVID-19 required admission to an intensive care unit: one at 30 weeks of gestation, after an emergency cesarean section;^19^ another case at 34 weeks of gestation that resulted in an emergency cesarean for a stillbirth;^31^ and other two cases at 37 weeks of gestation of women with high BMI (>35) and history of medical complications, admitted for labor induction.^12^ No maternal deaths from COVID-19 were published.

The 182 deliveries resulted in one stillbirth (intrauterine fetal death)^31^ and 185 live births (four twin pregnancies). There was one neonatal death in a preterm infant (34 completed weeks of gestation) from a pregnant woman with vaginal bleeding in the third trimester.^18^ Preterm birth occurred in 28.7% (45/157) among those with available information on gestational age. Approximately 16% of preterm births were spontaneous due to PROM^5, 18, 29, 31, 36^ or spontaneous onset of labor,^14^ but in most cases, it is unclear whether these were spontaneous or iatrogenic.

Although all breast milk samples from COVID-19 infected mothers have tested negative for the SARS-CoV-2 virus, most infants did not receive breast milk.

## Discussion

There is no evidence that the risk of infection with COVID-19 in pregnant women is greater than in the general population.^7, 37^ However, the incidence of infection in pregnant women is unknown, as screening tests were not generally used, except in the presence of symptoms. In a New York’s hospital that implemented universal SARS-CoV-2 testing in all pregnant women admitted for delivery, 15.4% of them were positive for SARS-CoV-2, but 87.9% were asymptomatic.^38^

Although most of the published cases confirm the absence of transmission of the SARS-CoV-2 virus antenatally or intrapartum, at least when the infection occurs in the third trimester of pregnancy,^5, 11, 13, 15, 18–26^ emerging evidence has suggested that vertical transmission is possible.^21, 24, 25, 27–29^ However, the evidence is still limited to a reduced number of reported cases, large variability in the type of biological material analyzed and the time of its collection. Even if vertical transmission occurred in the reported cases, the proportion would be low, below 5% of the published cases.

Regarding the effect of the SARS-CoV-2 virus on the fetus, no congenital malformation has been reported so far and the association of COVID-19 and fetal malformation seems unlikely considering the reduced risk of intrauterine infection.^37^

Higher risk of fetal distress and preterm births have been reported, but it is unclear if preterm birth occurs spontaneously (spontaneous onset of labor or following PROM) or is iatrogenic. The evidence related to the effect of COVID-19 on pregnant women is still limited. The clinical characteristics of COVID-19 were similar to those described in non-pregnant women, suggesting that the prognosis is not worse in pregnant women, although the number of cases studied is still reduced.^5, 18, 31–34^ A recent systematic review summarized the clinical manifestations of 108 pregnant women confirmed with COVID-19 and most of them presented fever (68%) and coughing (34%), and lymphocytopenia (59%) with elevated C-reactive protein (70%).^39^

The maternal and neonatal outcomes observed so far are quite different from the two most serious coronavirus-related previous epidemics.^7, 40–53^ The first also appeared in China, in 2002–03, and was characterized by severe respiratory infections caused by the Severe Acute Respiratory Syndrome-Coronavirus (SARS-CoV). The second occurred in 2012, initially in the Middle East, the Middle East Respiratory Syndrome - Coronavirus (MERS-CoV).^7, 40^ These epidemics have demonstrated the ability of coronavirus to cause serious complications during pregnancy,^43, 53^ with worse prognosis in pregnant women than non-pregnant women.^42, 54^

In the 2002 epidemic, 12 pregnant women were infected with SARS-CoV, with a fatality rate of 25%.^53^ Among the seven pregnant women infected in the first trimester, four had a miscarriage.^53^ Two of the five pregnant women infected during the second or third trimester had fetal growth restriction and four had a preterm delivery (one spontaneous; three induced by the maternal condition).^53^ In a review of the pregnancy outcomes of 11 women infected with MERS-CoV, seven pregnant women required admission to the intensive care unit and three died, of which only one had one comorbidity (asthma). Two fetal deaths occurred, and three of nine newborns were preterm.^43^

However, considering that SARS-CoV-2 has genetic homology and some clinical similarities to SARS-CoV and MERS-CoV, and the immunological and physiological changes that occur during pregnancy, such as in cell-mediated immunity or lung function, that affect both the susceptibility and the clinical severity of pneumonia, it is important to pay particular attention to the monitoring of pregnant women with COVID-19, because maternal and perinatal adverse outcomes are potentially relevant.^7, 40^ Although there were no maternal deaths described, possible maternal deaths were reported in social media, but without any robust evidence.^37^ Furthermore, one study reported two asymptomatic pregnant women at admission for delivery that rapidly evolved to severe COVID-19 disease requiring admission to an intensive care unit.^12^

Thus, it is essential to prevent the infection of COVID-19 and any other viral respiratory infection, as these infections represent an increased risk for the pregnant woman and for the pregnancy itself.^7, 55, 56^ It is therefore extremely important that pregnant women adopt preventive actions for COVID-19 with great intensity.^37^ For pregnant women with suspected or confirmed infection with SARS-CoV-2, recommendations for health professionals and services have already been published.^37, 55, 57–59^

Most women in this review had a cesarean section, many of them without a clear medical indication. The decision on the type of delivery in pregnant women with suspected or confirmed infection with COVID-19 should take into account the maternal and fetal clinical characteristics, as in normal practice, and not the diagnosis of COVID-19 infection per se. Thus, there is no obstetric contraindication to any mode of delivery, unless the pregnant woman’s clinical condition implies an emergent decision.^37^

In Portugal, we can estimate that about 80,000 pregnant women are exposed to the pandemic at different gestational ages, which represents an important challenge for individual and public health, to avoid infection in this population that should be considered at higher risk. According to the information reported by health professionals, there were eight cases of COVID-19 positive women delivered in Portugal by the 31st of March 2020. None of the first eight infants (all live births) born in Portugal tested positive for the SARS-CoV-2. Preterm birth occurred in 25% (2/8) of them and 5 (62.5%) resulted in cesarean section.

The limited scientific knowledge currently available makes it difficult to develop specific breastfeeding recommendations. There is not enough scientific evidence to unequivocally state that there is no possibility that mothers with COVID-19 can transmit the virus through breast milk.^55^ Therefore, recommendations should be based on the available data ^5 15, 16, 20–22, 28^ and the analogy with past circumstances and predictable costs and benefits. Breastfeeding is recognized as the best form of child feeding due to the countless benefits for both the mother and the newborn, including the protection against gastrointestinal and respiratory infections.^60^ Thus, considering the benefits of breastfeeding and the fact that the transmission of other respiratory viruses is insignificant through breast milk, there is no indication to stop breastfeeding. According to the recommendations of WHO/UNICEF^61^ and the Center for Disease Control and Prevention (CDC) of the United States,^55^ women with suspected or confirmed infection with COVID-19 can initiate or continue breastfeeding as long as clinical conditions permit. The CDC indicates that the decision to initiate or continue breastfeeding must be determined by the mother with COVID-19, together with family members and health professionals.^55^

Limitations of this systematic review should be acknowledged. Considerable heterogeneity was observed across the studies, which did not allow us to conduct a meta-analysis. On the other hand, we cannot guarantee that we were able to identify all the cases of pregnant women described in the literature. Possibly there are additional cases currently presented in other types of publications, such as reports. Also, considering the importance of summarizing all existing cases, we did not assess the quality of the studies included in this review. Several studies had missing outcome data and selective reporting bias could not be excluded. Additionally, there may be some cases which could be duplicated, namely the studies which did not describe clinical characteristics case by case.^30, 33, 34, 36^

## Conclusion

According to this review, fetal distress and preterm delivery seem to be more frequent among pregnant women with COVID-19. There is emerging evidence on possible vertical transmission (four positive results in the neonatal oral swabs for SARS-CoV-2 were reported and three newborns had high values of IgM antibodies), but the clinical relevance of the fetal infection is unclear. So far, there is no evidence that the SARS-CoV-2 virus is transmitted through breast milk. Maternal deaths were not reported and hospitalizations in intensive care were uncommon. Although the complications appear to be similar to those of non-pregnant women, services must be prepared to attend to complications, especially in pregnant women with comorbidities. Therefore, pregnant women and newborns should be considered particularly vulnerable populations regarding COVID-19 prevention and management strategies. Information, counseling and adequate monitoring are essential to prevent major adverse effects of SARS-CoV-2 infection during pregnancy.

## Data Availability

This is a systematic review.

## Acknowledgments

The authors are grateful to the health professionals who provided and confirmed the cases of pregnant women diagnosed with COVID-19 in Portugal: Alcides Pereira, Alexandra Almeida, Alexandrina Portela, Almerinda Pereira, Anselmo Costa, Gabriela Mimosa, Pedro Vieira da Silva, Rosalina Barroso, Teresa Rodrigues, Teresa Tomé.

## Ethics approval

This study is a systematic review based on published studies; therefore, ethical approval was not required.

## Authors’ contributions

Carina Rodrigues and Henrique Barros: Conceptualization, Original draft preparation. Inês Baía and Rosa Domingues: Methodology, Data curation, Writing-Reviewing, and Editing. All authors read and approved the final manuscript.

